# Validation of the PREDICT Breast Version 3.0 Prognostic Tool in US Breast Cancer Patients

**DOI:** 10.1101/2024.10.29.24316401

**Authors:** Yi-Wen Hsiao, Gordon C. Wishart, Paul D.P. Pharoah, Pei-Chen Peng

## Abstract

**Background:** PREDICT Breast v3 is the latest updated prognostication tool, developed from the breast cancer registry of approximately 35,000 women diagnosed between 2000 and 2018 in the United Kingdom. However, its performance in the United States (US) population is unknown. This study aims to validate PREDICT Breast v3 using newly released Surveillance, Epidemiology, and End Results (SEER) outcome data for US breast cancer patients and to address potential health disparities.

**Methods:** Over 860,000 female patients diagnosed between 2000 and 2018 with primary breast cancer and followed for at least 10 years were selected from the SEER database. Predicted and observed 10- and 15-year breast cancer-specific survival outcomes were compared for the overall cohort, stratified by estrogen receptor (ER) status, and predefined subgroups. Discriminatory accuracy was determined through the area under the receiver-operator curves (AUC).

**Results:** PREDICT Breast v3 demonstrated good calibration and discrimination for long-term breast cancer-specific mortality. It provided accurate mortality estimates (within a ±10% error range) across the entire US population for 10-year (−8% in ER-positive and 4% in ER-negative patients) and 15-year (−3 % in ER-positive and 5% in ER-negative patients) all-cause mortality, for both ER statuses. The model also showed good performance for 10- and 15-year all-cause mortality across the U.S. population, with AUC of 0.769 and 0.793 for ER-positive breast cancer as well as AUC of 0.738 and 0.746 for ER-negative breast cancer. However, recalibration is needed for specific groups, such as non-Hispanic Asian and non-Hispanic Black patients with ER-negative status.

**Conclusions:** PREDICT v3 accurately predicts 10- and 15-year overall survival in contemporary US breast cancer patients. Future work should focus on promoting equitable care by addressing disparities that are observed in predictive tools.

## 1 Background

Breast cancer is the most common type of cancer diagnosed among women worldwide, with around 2.3 million new cases diagnosed in 2022 ^1^. It also has the highest incidence rates and the second-largest mortality rates among women in the US, regardless of race or ethnicity ^2^: in the United States, a total of 310,720 new female breast cases and 42,250 breast cancer-related deaths among women are estimated in 2024 ^2^. A key decision for women with a new diagnosis of breast cancer, made in discussion with their clinicians, is whether to undergo a course of systemic treatment.

Adjuvant systemic treatment after surgery for early-stage breast cancer patients is aimed at reducing the risk of recurrence and mortality ^3^. Accurate estimates of survival and the benefit of such treatment in early-stage breast cancer ensures that potentially harmful treatment is targeted to those most likely to benefit. These estimates can help oncologists to support optimal clinical decision making which reduces the side effects and maintain the quality of life for breast cancer patients ^4^. Prediction models such as PREDICT Breast ^5^, Adjuvant! Online ^6^, and CancerMath ^7^ were developed to help decide which adjuvant systemic therapy is most suitable for the patient depending on the patient and tumor characteristics, including tumor size, node status, hormone receptor status, and other factors ^8^. Adjuvant! Online is no longer available and CancerMath has not been updated since it was first released. PREDICT Breast has been regularly modified and updated since it was released in 2011; PREDICT Breast v3 (breast.v3.predict.cam) ^9^, released in May 2024, is the most recent version.

PREDICT Breast v1 and v2 have been validated in breast cancer cases from multiple countries, including UK ^10,11^, Canada ^12^, Malaysia ^13^, the Netherlands ^14,14,15^, Japan ^16^, Indian ^17^, Spain ^18^ and New Zealand ^19^ and the USA ^20,20^. However, PREDICT Breast v3, has only been validated in breast cancer cases from the UK – the population used to develop the model. It is important that this version should be validated in other populations including the USA and, given the diversity of the population in the USA, it is important to evaluate the performance of PREDICT Breast v3 in the different racial groups within the USA. Thus, this study aims to address this gap by conducting an external validation of PREDICT Breast v3 using the latest release of the Surveillance, Epidemiology, and End Results (SEER) data. This validation will assess the model’s accuracy in predicting patient outcomes across diverse populations of breast cancer patients in the United States. We have also compared the performance of PREDICT Breast v3 with that of PREDICT v2.2 and CancerMath.

## 2 Methods

### 2.1 Study population

The study population was from the SEER Research Plus data (2000-2018; November 2023 Submission) ^21^. SEER is a comprehensive cancer registry program in the US that collects information about cancer patients from multiple cancer registries across the country. In total, 1,291,324 breast cancer cases were recorded in this latest release. The breast cancer registry captures information on patient demographics, the tumor site, time since initial cancer diagnosis, tumor histology and tumor behavior.

In this study, women aged 25 to 84 diagnosed with primary breast cancer in 2000 through 2018 were included. Patients with distant metastasis at the time of diagnosis, tumor size exceeding 500 mm or more than 50 positive lymph nodes were excluded, as were those with missing information necessary for PREDICT Breast prognostic score calculations. We also excluded the patients with missing data on survival time or cause of death. The final cohort comprised 628,753 female breast cancer patients: 62,402 Hispanic (All Races), 51,136 Non-Hispanic Asian or Pacific Islander, 57,039 Non-Hispanic Black and 453,297 Non-Hispanic White.

### 2.2 SEER prognostic variables used in the PREDICT Breast model

The minimum set of input variables for PREDICT Breast v3 are patient demographics (diagnosis year and age at diagnosis), tumor characteristics (tumor size, histologic grade, number of positive lymph nodes, estrogen receptor status), and treatment types (radiation therapy, adjuvant hormone therapy, adjuvant chemotherapy, trastuzumab and bisphosphonates). Adjuvant chemotherapy is either classified as standard anthracycline based or high-dose anthracycline/taxane based. Optional variables are mode of detection (clinical or screen detected) and tumor HER2 status, progesterone receptor status and KI67 status. KI67 data are not available in SEER and so KI67 status was set to unknown for all cases. Mode of detection is also unavailable and was assumed to be screening for 20% of cases aged 45-69 and clinically detected for all others.

SEER also provides information on race and ethnicity encoded as non-Hispanic White, non-Hispanic Black, non-Hispanic American Indian or Alaska Native, non-Hispanic Asian or Pacific Islander, and Hispanic. Each variable is either quantitative or categorical, as detailed in Appendix 1.

Treatment data in SEER are limited to indicator variables for radiotherapy and adjuvant chemotherapy with no data on adjuvant hormone therapy, trastuzumab or bisphosphonate therapy. We assumed that: 1) all cases diagnosed under age 65 who received chemotherapy had a high-dose anthracycline/taxane based and all those 65 and over had a standard anthracycline based regimen; 2) all ER-positive patients received hormone therapy, (3) those diagnosed after 2000 with HER2-positive cancer were assumed to have been prescribed trastuzumab; and (4) no patients received bisphosphonate treatment.

### 2.3 Calculating the PREDICT breast v3 predicted survival probabilities

Predicted all-cause mortality for PREDICT Breast v3 was calculated using a custom script based on the model described in Grootes at al ^9^. PREDICT takes the form of a competing risk Cox survival model, with fractional polynomial baseline cumulative hazards. The competing risks are breast cancer mortality and other mortality.

The estimated survival from breast cancer at t years after surgery for each patient is given by

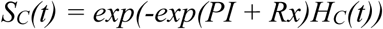

PI is the breast cancer prognostic index (log hazard ratio) given by

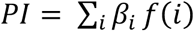

for prognostic factors 1…*i* where *β_i_* and *f(i)* are the log relative hazards and the function of *i* respectively.

Rx is the effect of treatment (log hazard ratio) given by

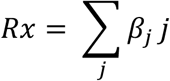

for treatments 1..*j,* where *β_j_* is the log relative hazard for treatment *j*.

H_C_(t) is the baseline hazard for breast cancer mortality.

The estimated survival from other causes is given by

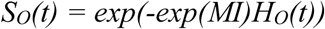

MI is the mortality index

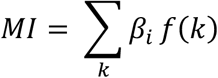

for other mortality prognostic factors 1…*k* where *β_k_* and *f(k)* are the coefficients and the function of *k*.

The <u>all-cause survival function</u> at time *t* assumes independent, competing risks from breast and other mortality and is given by

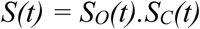

Thus, predicted all-cause mortality at time t is given by

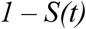

The baseline hazard functions, and coefficients and functions for all breast cancer and non-breast cancer risk factors were taken from Grootes et al ^9^.

Predicted all-cause mortality for PREDICT Breast v2.2 was calculated using the *nhs.predict* R package ^15^. The predicted all-cause mortality for CancerMath was calculated using a custom R script derived from the JavaScript extracted from the online tool. The output of the R script was verified comparing them with those generated by the online tool for a small set of cases.

### 2.4 Predictive model performance

Model performance was evaluated using calibration, goodness-of-fit and discrimination. Model calibration is given by the ratio of the observed number of events divided by the number of events predicted by the model. Goodness-of-fit was assessed graphically by plotting the observed number of deaths against the predicted number of deaths within quintiles of risk. Model discrimination was evaluated by calculating the area under the receiver-operator characteristic curve (AUC) which measures the probability that the predicted mortality score for a randomly selected patient who died will be higher than that for a randomly selected patient who survived. An AUC value ranges between 0.5 to 1, with a higher AUC indicating a better model in identifying patients with a worse survival. AUC statistics were calculated separately for different ER status and different populations.

All analyses were conducted using the R software ^22^ implemented in the R Studio version 4.3.3 ^23^ and the packages *survival* ^24^, *pROC* ^25^ and *tidyverse* ^26^.

## 3 RESULTS

### 3.1 Demographic and clinical characteristics of breast cancer patients in SEER

The study population included 628,753 women diagnosed with breast cancer during 2000 to 2018 in the SEER cancer registry, with 712,233 having ER–positive status and 148,751 having ER–negative status. Of these, 413,280 had a minimum of 10 years follow up (83,084 ER-positive and 330,196 ER-negative) and 220,022 had a minimum of 15 years follow up (172,307 ER+ and 47,715 ER-). Table 1 summarizes patients demographics, tumor characteristics, and treatment types, stratified by ER status.

**Table 1.**
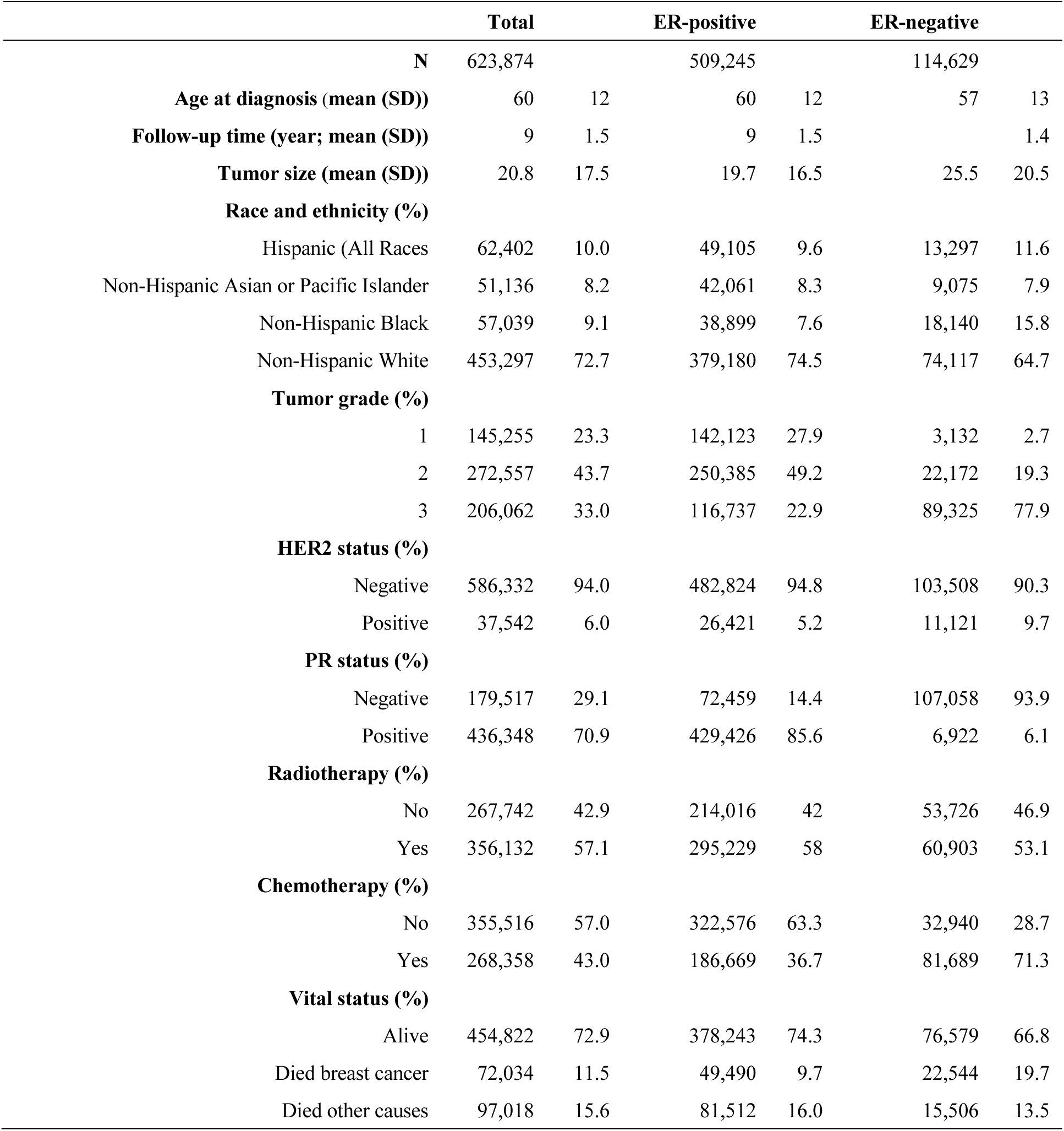
Breast cancer patients’ demographics, tumor characteristics, and treatment types in SEER, stratified by estrogen receptor status. Characteristics were summarized using the proportions for categorical variables and mean (standard derivation, SD) for continuous variables.

|  | Total |  | ER-positive |  | ER-negative |  |
| --- | --- | --- | --- | --- | --- | --- |
| N | 623,874 |  | 509,245 |  | 114,629 |  |
| Age at diagnosis (mean (SD)) | 60 | 12 | 60 | 12 | 57 | 13 |
| Follow-up time (year; mean (SD)) | 9 | 1.5 | 9 | 1.5 |  | 1.4 |
| Tumor size (mean (SD)) | 20.8 | 17.5 | 19.7 | 16.5 | 25.5 | 20.5 |
| **Race and ethnicity (%)** |  |  |  |  |  |  |
| Hispanic (All Races | 62,402 | 10.0 | 49,105 | 9.6 | 13,297 | 11.6 |
| Non-Hispanic Asian or Pacific Islander | 51,136 | 8.2 | 42,061 | 8.3 | 9,075 | 7.9 |
| Non-Hispanic Black | 57,039 | 9.1 | 38,899 | 7.6 | 18,140 | 15.8 |
| Non-Hispanic White | 453,297 | 72.7 | 379,180 | 74.5 | 74,117 | 64.7 |
| **Tumor grade (%)** |  |  |  |  |  |  |
| 1 | 145,255 | 23.3 | 142,123 | 27.9 | 3,132 | 2.7 |
| 2 | 272,557 | 43.7 | 250,385 | 49.2 | 22,172 | 19.3 |
| 3 | 206,062 | 33.0 | 116,737 | 22.9 | 89,325 | 77.9 |
| **HER2 status (%)** |  |  |  |  |  |  |
| Negative | 586,332 | 94.0 | 482,824 | 94.8 | 103,508 | 90.3 |
| Positive | 37,542 | 6.0 | 26,421 | 5.2 | 11,121 | 9.7 |
| **PR status (%)** |  |  |  |  |  |  |
| Negative | 179,517 | 29.1 | 72,459 | 14.4 | 107,058 | 93.9 |
| Positive | 436,348 | 70.9 | 429,426 | 85.6 | 6,922 | 6.1 |
| **Radiotherapy (%)** |  |  |  |  |  |  |
| No | 267,742 | 42.9 | 214,016 | 42 | 53,726 | 46.9 |
| Yes | 356,132 | 57.1 | 295,229 | 58 | 60,903 | 53.1 |
| **Chemotherapy (%)** |  |  |  |  |  |  |
| No | 355,516 | 57.0 | 322,576 | 63.3 | 32,940 | 28.7 |
| Yes | 268,358 | 43.0 | 186,669 | 36.7 | 81,689 | 71.3 |
| **Vital status (%)** |  |  |  |  |  |  |
| Alive | 454,822 | 72.9 | 378,243 | 74.3 | 76,579 | 66.8 |
| Died breast cancer | 72,034 | 11.5 | 49,490 | 9.7 | 22,544 | 19.7 |
| Died other causes | 97,018 | 15.6 | 81,512 | 16.0 | 15,506 | 13.5 |

### 3.2 Calibration

Overall, PREDICT Breast v3 was well-calibrated. The predicted number of deaths at 10 years was within 10% of the observed deaths in patients with ER-positive cancers (68,114 predicted/74,326 observed) and in patients with ER-negative cancer (26,244 predicted/25,190 observed). At 15-years the predictions were within 5% (ER-positive patients 61,770 predicted/63,718 observed; ER-negative patients 20,191 predicted/19,217 observed). The observed and predicted number of deaths from all-causes at 10 and 15 years stratified by patient demographics, tumor characteristics, and treatment are shown in Table 2 for ER-positive patients and Table 3 for ER-negative patients. In most subgroups the observed and predicted number of deaths were within 10%. However, calibration was poor in non-Hispanic Asians with ER-negative cancer, with PREDICT Breast v3 over-predicting the number of deaths at 10- and 15-years by more than 30%. Similarly, calibration in non-Hispanic black women with ER-positive breast cancer was poor with PREDICT Breast v3 under-predicting the number of deaths by 20% or more at 10- and 15-years. The observed and predicted numbers of deaths from breast cancer are shown in Appendix 2 for ER-positive breast cancer and in Appendix 3 for ER-negative breast cancer, with deaths from other causes shown in Appendix 4 for ER-positive breast cancer and in Appendix 5 for ER-negative breast cancer. In general, breast cancer specific mortality tended to be over-estimated whereas mortality from other causes tended to be under-estimated.

**Table 2.** Cumulative observed and predicted all-cause mortality at 10 and 15 years follow up for ER-positive patients.

| Characteristics | 10-year |  |  |  |  |  | 15-year |  |  |  |  |
| --- | --- | --- | --- | --- | --- | --- | --- | --- | --- | --- | --- |
|  | N | O | P | D | % |  | N | O | P | D | % |
| **Total** | 327,873 | 74,326 | 68,114 | -6,212 | -8.4 |  | 171,194 | 63,378 | 61,770 | -1,608 | -2.5 |
| **Age at diagnosis** |  |  |  |  |  |  |  |  |  |  |  |
| <36y | 5,925 | 1,214 | 1,017 | -197 | -16 |  | 3,108 | 932 | 877 | -55 | -5.9 |
| 36-40 y | 11,900 | 1,759 | 1,566 | -193 | -11 |  | 6,481 | 1,465 | 1,456 | -9 | -0.6 |
| 41-50 y | 63,908 | 7,053 | 6,894 | -159 | -2.3 |  | 34,024 | 5,925 | 6,598 | 673 | 11 |
| 51-60 y | 84,863 | 11,570 | 10,465 | -1,105 | -10 |  | 44,634 | 9,962 | 10,087 | 125 | 1.3 |
| >60 y | 161,277 | 52,730 | 48,170 | -4,560 | -8.6 |  | 82,947 | 45,094 | 42,752 | -2,342 | -5.2 |
| **Race and ethnicity** |  |  |  |  |  |  |  |  |  |  |  |
| Hispanic (All Races) | 28,532 | 5,865 | 5,479 | -386 | -6.6 |  | 13,381 | 4,438 | 4,470 | 32 | 0.7 |
| Non-Hispanic Asian or Pacific Islander | 24,937 | 3,862 | 4,406 | 544 | 14 |  | 12,043 | 3,188 | 3,729 | 541 | 17 |
| Non-Hispanic Black | 23,508 | 6,978 | 4,912 | -2,066 | -30 |  | 11,185 | 5,011 | 4,019 | -992 | -20 |
| Non-Hispanic White | 250,896 | 57,621 | 53,315 | -4,306 | -7.5 |  | 134,585 | 50,741 | 49,552 | -1,189 | -2.3 |
| **Tumor size** |  |  |  |  |  |  |  |  |  |  |  |
| 0-10 mm | 93,252 | 15,047 | 14,306 | -741 | -4.9 |  | 47,998 | 14,396 | 13,903 | -493 | -3.4 |
| 11-20 mm | 130,997 | 26,320 | 24,226 | -2,094 | -8.0 |  | 70,300 | 24,086 | 23,288 | -798 | -3.3 |
| 21-50 mm | 88,621 | 26,684 | 23,785 | -2,899 | -11 |  | 45,588 | 20,765 | 20,276 | -489 | -2.4 |
| >50 mm | 15,003 | 6,275 | 5,795 | -480 | -7.6 |  | 7,308 | 4,131 | 4,302 | 171 | 4.1 |
| **Tumor nodes** |  |  |  |  |  |  |  |  |  |  |  |
| 0 | 222,802 | 42,409 | 39,008 | -3,401 | -8.0 |  | 114,026 | 38,060 | 36,521 | -1,539 | -4.0 |
| 1 | 45,003 | 10,333 | 9,218 | -1,115 | -11 |  | 24,062 | 8,708 | 8,490 | -218 | -2.5 |
| 2-4 | 35,956 | 10,221 | 9,382 | -839 | -8.2 |  | 19,786 | 8,458 | 8,442 | -16 | -0.2 |
| 5-9 | 14,881 | 6,213 | 5,607 | -606 | -10 |  | 8,232 | 4,635 | 4,660 | 25 | 0.5 |
| 10+ | 9,231 | 5,150 | 4,897 | -253 | -4.9 |  | 5,088 | 3,517 | 3,658 | 141 | 4.0 |
| **Tumor grade** |  |  |  |  |  |  |  |  |  |  |  |
| 1 | 89,562 | 16,134 | 14,074 | -2,060 | -13 |  | 45,032 | 14,671 | 13,271 | -1,400 | -10 |
| 2 | 159,627 | 35,749 | 31,758 | -3,991 | -11 |  | 83,313 | 30,726 | 29,037 | -1,689 | -5.5 |
| 3 | 78,684 | 22,443 | 22,281 | -162 | -0.7 |  | 42,849 | 17,981 | 19,462 | 1,481 | 8.2 |
| **HER2 status** |  |  |  |  |  |  |  |  |  |  |  |
| negative | 52,502 | 11,420 | 9,347 | -2,073 | -18 |  | - | - | - | - | - |
| positive | 6,577 | 1,316 | 1,038 | -278 | -21 |  | - | - | - | - | - |
| **PR status** |  |  |  |  |  |  |  |  |  |  |  |
| negative | 49,184 | 13,766 | 11,627 | -2,139 | -16 |  | 26,729 | 11,254 | 10,679 | -575 | -5.1 |
| positive | 271,694 | 58,946 | 54,979 | -3,967 | -6.7 |  | 139,421 | 50,236 | 49,260 | -976 | -1.9 |
| **Chemotherapy** | 124,751 | 25,039 | 23,260 | -1,779 | -7.1 |  | 66,451 | 20,405 | 20,752 | 347 | 1.7 |
| **Radiotherapy** | 186,235 | 36,103 | 35,904 | -199 | -0.6 |  | 97,582 | 31,920 | 32,806 | 886 | 2.8 |

N: number of cases; O: observed number of deaths; P: predicted number of deaths; D: difference of the number of deaths between predicted and observed; %: percentage of the difference of the number of deaths between predicted and observed.

**Table 3.** Cumulative observed and predicted all-cause mortality at 10 and 15 years follow up for ER-negative patients.

| Characteristics | 10-year |  |  |  |  | 15-year |  |  |  |  |
| --- | --- | --- | --- | --- | --- | --- | --- | --- | --- | --- |
|  | N | O | P | D | % | N | O | P | D | % |
| **Total** | 82,450 | 25,190 | 26,244 | 1,054 | 4.2 | 47,374 | 19,217 | 20,191 | 974 | 5.1 |
| **Age at diagnosis** |  |  |  |  |  |  |  |  |  |  |
| <36y | 3,668 | 902 | 1,263 | 361 | 40 | 2,239 | 615 | 935 | 320 | 52 |
| 36-40 y | 5,369 | 1,251 | 1,507 | 256 | 20 | 3,316 | 946 | 1,155 | 209 | 22 |
| 41-50 y | 19,165 | 4,470 | 4,670 | 200 | 4.5 | 11,530 | 3,351 | 3,641 | 290 | 8.7 |
| 51-60 y | 23,735 | 5,829 | 5,895 | 66 | 1.1 | 13,592 | 4,376 | 4,582 | 206 | 4.7 |
| >60 y | 30,513 | 12,738 | 12,909 | 171 | 1.3 | 16,697 | 9,929 | 9,879 | -50 | -0.5 |
| **Race and ethnicity** |  |  |  |  |  |  |  |  |  |  |
| Hispanic (All Races) | 8,962 | 2,588 | 2,818 | 230 | 8.9 | 4,801 | 1,842 | 2,005 | 163 | 8.8 |
| Non-Hispanic Asian or Pacific Islander | 6,067 | 1,325 | 1,821 | 496 | 37 | 3,271 | 999 | 1,319 | 320 | 32 |
| Non-Hispanic Black | 12,548 | 4,513 | 4,050 | -463 | -10 | 6,903 | 3,123 | 2,929 | -194 | -6.2 |
| Non-Hispanic White | 54,873 | 16,764 | 17,556 | 792 | 4.7 | 32,399 | 13,253 | 13,938 | 685 | 5.2 |
| **Tumor size** |  |  |  |  |  |  |  |  |  |  |
| 0-10 mm | 14,252 | 2,599 | 2,558 | -41 | -1.6 | 7,967 | 2,419 | 2,261 | -158 | -6.5 |
| 11-20 mm | 27,611 | 6,949 | 7,059 | 110 | 1.6 | 16,354 | 5,771 | 5,922 | 151 | 2.6 |
| 21-50 mm | 33,872 | 12,048 | 12,635 | 587 | 4.9 | 19,399 | 8,776 | 9,426 | 650 | 7.4 |
| >50 mm | 6,715 | 3,594 | 3,991 | 397 | 11 | 3,654 | 2,251 | 2,583 | 332 | 15 |
| **Tumor nodes** |  |  |  |  |  |  |  |  |  |  |
| 0 | 52,840 | 12,317 | 12,603 | 286 | 2.3 | 29,602 | 10,090 | 10,170 | 80 | 0.8 |
| 1 | 10,953 | 3,598 | 3,597 | -1 | 0.0 | 6,369 | 2,634 | 2,750 | 116 | 4.4 |
| 2-4 | 10,102 | 4,120 | 4,409 | 289 | 7.0 | 6,123 | 2,994 | 3,306 | 312 | 10 |
| 5-9 | 4,866 | 2,659 | 2,878 | 219 | 8.2 | 3,005 | 1,844 | 2,084 | 240 | 13 |
| 10+ | 3,689 | 2,496 | 2,756 | 260 | 10 | 2,275 | 1,655 | 1,880 | 225 | 14 |
| **Tumor grade** |  |  |  |  |  |  |  |  |  |  |
| 1 | 2,475 | 531 | 539 | 8 | 1.5 | 1,573 | 526 | 520 | -6 | -1.1 |
| 2 | 15,955 | 4,726 | 4,334 | -392 | -8.3 | 9,315 | 3,866 | 3,605 | -261 | -6.8 |
| 3 | 64,020 | 19,933 | 21,372 | 1,439 | 7.2 | 36,486 | 14,825 | 16,067 | 1,242 | 8.4 |
| **HER2 status** |  |  |  |  |  |  |  |  |  |  |
| negative | 8,849 | 2,808 | 2,409 | -399 | -14 | - | - | - | - | - |
| positive | 2,932 | 700 | 685 | -15 | -2.1 | - | - | - | - | - |
| **PR status** |  |  |  |  |  |  |  |  |  |  |
| negative | 76,941 | 23,729 | 24,615 | 886 | 3.7 | 43,808 | 17,928 | 18,803 | 875 | 4.9 |
| positive | 4,903 | 1,292 | 1,434 | 142 | 11 | 3,116 | 1,105 | 1,199 | 94 | 8.5 |
| **Chemotherapy** | 56,687 | 15,971 | 17,012 | 1,041 | 6.5 | 31,552 | 11,665 | 12,624 | 959 | 8.2 |
| **Radiotherapy** | 43,083 | 12,014 | 12,880 | 866 | 7.2 | 25,196 | 9,259 | 10,093 | 834 | 9.0 |

N: number of cases; O: observed number of deaths; P: predicted number of deaths; D: difference of the number of deaths between predicted and observed; %: percentage of the difference of the number of deaths between predicted and observed.

Results of calibration at 5-years follow-up for all-cause, breast cancer-specific and other-cause mortalities are shown in Appendix 6 for ER-positive breast cancer and in Appendix 7 for ER-negative breast cancer. PREDICT breast v3 tended to under-estimate both breast cancer and other deaths at five years with a more substantial mis-calibration for ER-positive patients.

### 3.3 Goodness-of-fit

The comparison between predicted and observed all-cause mortality by quintiles of predicted risk is shown in Figure 1. Overall, PREDICT Breast demonstrated good calibration across most quartiles. Mis-calibration was greatest in patients at highest risk. Goodness-of-fit plots for breast cancer-specific and other causes of mortality at 10 and 15 years are shown in Appendix 8.

**Figure 1.**
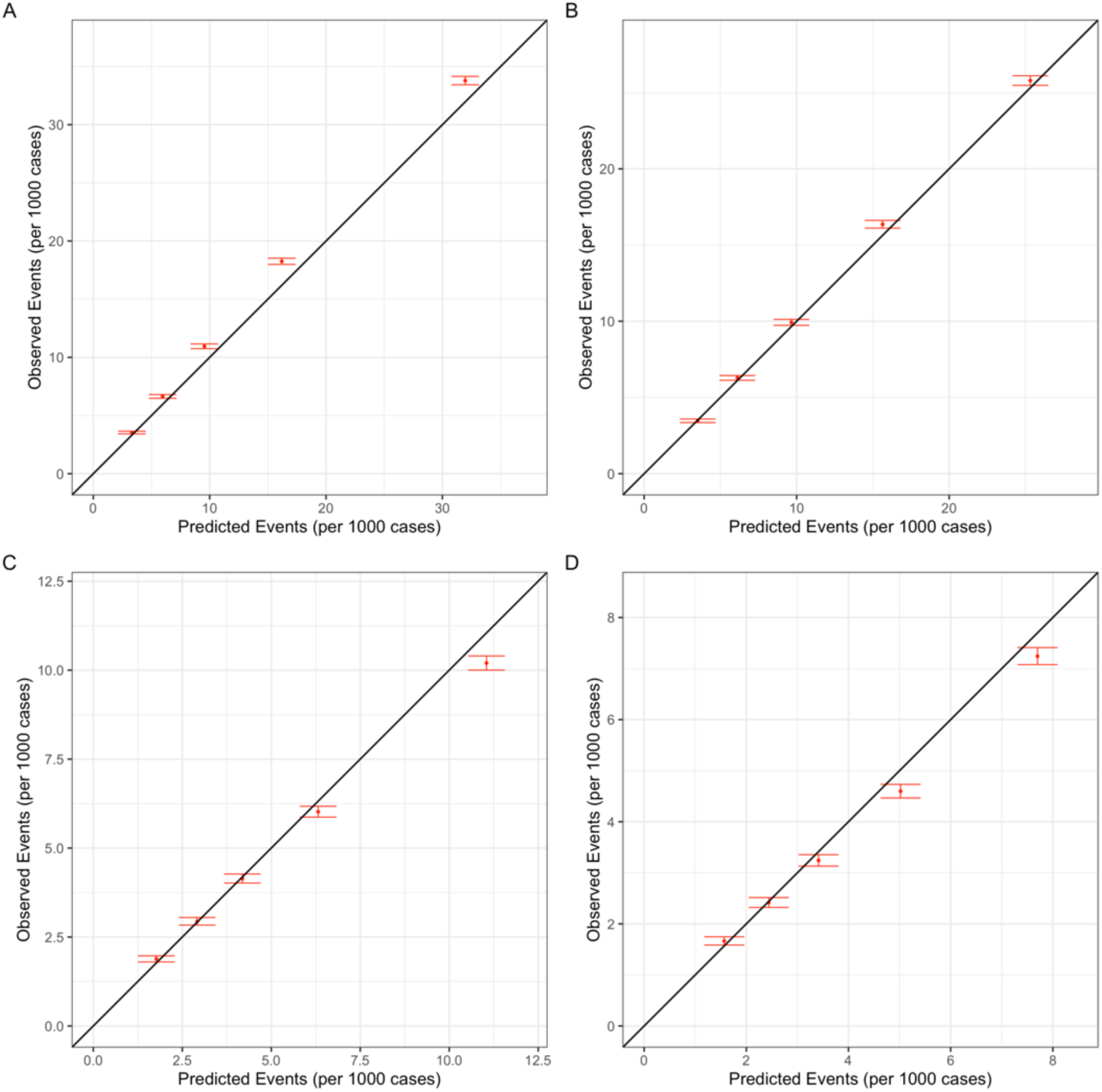
Calibration plot for all-cause mortality by follow up time and tumor ER-status. (A) 10-year, ER-positive, (B) 15-year, ER-positive, (C) 10-year, ER-negative, (D) 15-year, ER-negative.

Goodness-of-fit plots for all-cause mortality, breast cancer-specific mortality and other causes of mortality are shown in Appendix 9.

### 3.4 Discrimination

Overall model discrimination was very good in women with both ER-positive breast cancer (AUCs of 0.769 for 10-year follow-up and 0.793 for 15-year follow-up) and ER-negative breast cancer (AUCs of 0.738 for 10-year follow-up and 0.746 for 15-year follow-up) (Table 4). There was little difference in discrimination by race. Discrimination was slightly better for other cause mortality than for breast cancer specific mortality is shown in Appendix 10. AUCs for 5-year mortality were similar (Appendix 11).

**Table 4.** Model discrimination (area under receiver operator characteristic curve) for 10-year and 15-year all-cause mortality by race and tumor ER-status.

| Race | ER-positive |  | ER-negative |  |
| --- | --- | --- | --- | --- |
|  | 10-year | 15-year | 10-year | 15-year |
| **Total** | 0.769 | 0.793 | 0.738 | 0.746 |
| Non-Hispanic White | 0.774 | 0.800 | 0.725 | 0.727 |
| Non-Hispanic Asian | 0.758 | 0.764 | 0.725 | 0.756 |
| Hispanic (All Races) | 0.755 | 0.767 | 0.746 | 0.717 |
| Non-Hispanic Black | 0.737 | 0.754 | 0.742 | 0.737 |

### 3.5 Comparison of performance of PREDICT breast v3 with v2.2 and CancerMath

Calibration and discrimination for all three models for all-cause mortality at 10 and 15 years are shown in Table 5. Both PREDICT v2.2 and CancerMath substantially over predicted the number of deaths with calibration being particularly poor for CancerMath. Discrimination was good for all three models, with PREDICT v3 slightly outperforming the other two models.

**Table 5.** Performance comparison of other breast cancer prognostication tools with PREDICT v3 in the US population, in terms of the 10-year and 15-year all-cause mortality, stratified by ER status.

|  | ER-positive |  | ER-negative |  |
| --- | --- | --- | --- | --- |
|  | Calibration (%) | Discrimination | Calibration (%) | Discrimination |
| **10-year** |  |  |  |  |
| PREDICT Breast v3 | -8.4 | 0.769 | 4.2 | 0.738 |
| PREDICT Breast v2 | 31 | 0.758 | 39 | 0.732 |
| CancerMath | 83 | 0.740 | 32 | 0.710 |
| **15-year** |  |  |  |  |
| PREDICT Breast v3 | -2.5 | 0.793 | 5.1 | 0.746 |
| PREDICT Breast v2 | 19 | 0.774 | 23 | 0.741 |
| CancerMath | 112 | 0.740 | 64 | 0.710 |

## 4 DISCUSSION

This study is the first validation of PREDICT Breast v3 in a non-UK population. Overall, the model performed well with good calibration and discrimination at 10 and 15 years for ER-positive and ER-negative patients. The overall performance was similar to that in a large series of patients in the United Kingdom ^9^. Discrimination was generally very good in all populations for both ER-negative and ER-positive patients, but calibration was poorer in specific populations. In particular, the model over estimated mortality in non-Hispanic Asian patients with ER-negative disease and under-estimated mortality in non-Hispanic black patients with ER-positive disease. The latter finding is consistent with findings from a validation of PREDICT breast v2 in the US population ^20^.

The primary purpose of PREDICT breast is to provide estimates of the absolute survival benefit associated with adjuvant therapies to aid shared decision making between patients and their oncologists. Model performance indicates that PREDICT breast v3 is sufficiently accurate in the US non-Hispanic white population for it to be incorporated into the routine practice of oncologists. However, the model is likely to over-estimate the benefits of adjuvant therapy in non-Hispanic Asian patients with ER-negative disease and under-estimate the benefits of adjuvant therapy in the non-Hispanic black patients with ER-positive disease. The under/over-estimates are by about one third at 10 years, and this should be taken into account when using the model for decision making in these populations.

There are two main components to the PREDICT breast model. The first is the baseline hazard and the second is the set of coefficients (log hazard ratios) for each prognostic factor in the model. Poor calibration is primarily dependent on misspecification of the baseline hazard, whereas discrimination depends on the set of coefficients. Given that discrimination was good across all population groups, completely refitting a model to generate different sets of population specific coefficients is unlikely to improve the fit of the model substantially. However, improvements in calibration could easily be achieved by simply modifying the baseline hazard to be population specific.

A further limitation is a limitation of the PREDICT breast v3 model itself. There are many markers that have been shown to be prognostic in addition to the variables included in the model. Of particular note are tumor gene expression profiles or genomic risk scores (GRS) such as EndoPredict ^27^, Mammaprint ^28^, and OncotypeDx ^29^. While there are many published ‘validation’ studies of GRS, there has only been one study to evaluate the benefit of adding GRS to standard clinical variables as measured by change in discrimination or reclassification ^30^; in this study, adding GRS to PREDICT breast v2 had a small effect on the discrimination of the model and reclassification was limited. It seems unlikely that adding GRS to PREDICT breast v3 would make much difference to the performance.

We have shown that PREDICT breast v3 works well for the majority of breast cancer patients in the USA. Future work will involve evaluating the benefit of adding GRS to the model and modification of the model to ensure that performance is good in all ancestries to reflect the diverse ancestries of the population.

## Data Availability

The data used in this study are available from the National Cancer Institute SEER program, at https://seer.cancer.gov/.

## ACKNOWLEDGEMENT

This research was supported by the Cedars-Sinai Cancer Center through the 2024 Cancer Prevention and Control Program Research Developmental Funds Award.

## DATA AND CODE AVAILABILITY

The data used in this study are available from the National Cancer Institute SEER program, at https://seer.cancer.gov/. The R script utilized for data analysis can be accessed on GitHub at https://github.com/pengpclab/PREDICTv3.

**Appendix 1.**
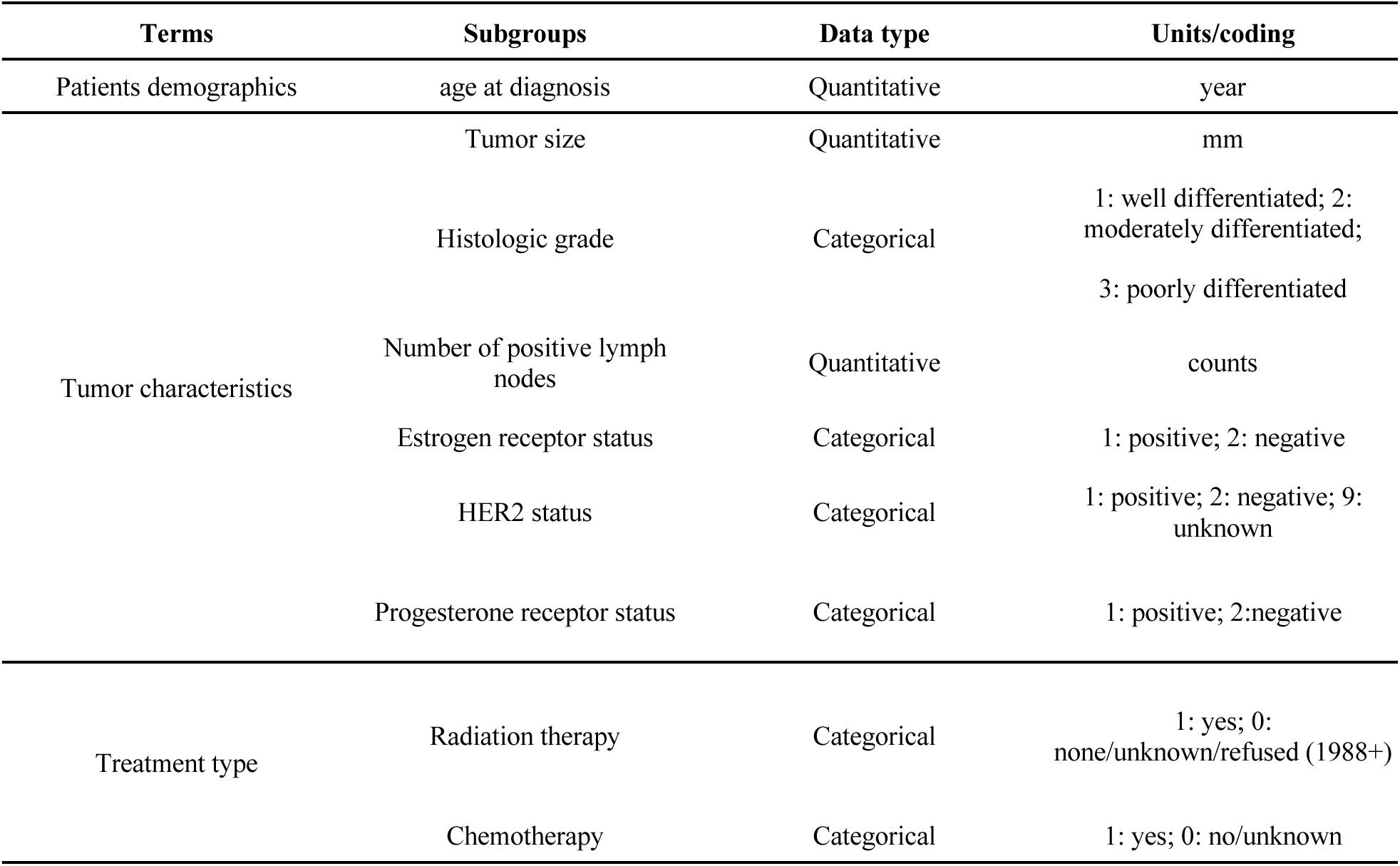
Summary of standard input variables used in the PREDICT Breast model.

**Appendix 2.** Cumulative observed and predicted breast cancer-specific mortality at 10 and 15 years follow up for ER-positive patients.

| Characteristics | 10-year |  |  |  |  | 15-year |  |  |  |  |
| --- | --- | --- | --- | --- | --- | --- | --- | --- | --- | --- |
|  | N | O | P | D | % | N | O | P | D | % |
| **Total** | 327,873 | 30,932 | 31,129 | 197 | 0.6 | 171,194 | 23,221 | 26,000 | 2,779 | 12 |
| **Age at diagnosis** |  |  |  |  |  |  |  |  |  |  |
| <36y | 5,925 | 1,121 | 941 | -180 | -16 | 3,108 | 838 | 785 | -53 | -6.3 |
| 36-40 y | 11,900 | 1,552 | 1,390 | -162 | -10 | 6,481 | 1,263 | 1,238 | -25 | -2 |
| 41-50 y | 63,908 | 5,512 | 5,548 | 36 | 0.7 | 34,024 | 4,402 | 4,978 | 576 | 13 |
| 51-60 y | 84,863 | 7,058 | 6,901 | -157 | -2.2 | 44,634 | 5,518 | 5,959 | 441 | 8 |
| >60 y | 161,277 | 15,689 | 16,348 | 659 | 4.2 | 82,947 | 11,200 | 13,039 | 1,839 | 16 |
| **Race and ethnicity** |  |  |  |  |  |  |  |  |  |  |
| Hispanic (All Races) | 28,532 | 3,152 | 3,058 | -94 | -3 | 13,381 | 2,135 | 2,315 | 180 | 8.4 |
| Non-Hispanic Asian or Pacific Islander | 24,937 | 1,971 | 2,293 | 322 | 16 | 12,043 | 1,477 | 1,814 | 337 | 23 |
| Non-Hispanic Black | 23,508 | 3,552 | 2,730 | -822 | -23 | 11,185 | 2,319 | 2,099 | -220 | -9.5 |
| Non-Hispanic White | 250,896 | 22,257 | 23,047 | 790 | 3.6 | 134,585 | 17,290 | 19,771 | 2,481 | 14 |
| **Tumor size** |  |  |  |  |  |  |  |  |  |  |
| 0-10 mm | 93,252 | 3,078 | 3,063 | -15 | -0.5 | 47,998 | 2,624 | 2,864 | 240 | 9.2 |
| 11-20 mm | 130,997 | 8,755 | 9,212 | 457 | 5.2 | 70,300 | 7,325 | 8,406 | 1,081 | 15 |
| 21-50 mm | 88,621 | 14,705 | 14,466 | -239 | -1.6 | 45,588 | 10,569 | 11,649 | 1,080 | 10 |
| >50 mm | 15,003 | 4,394 | 4,387 | -7 | -0.2 | 7,308 | 2,703 | 3,080 | 377 | 14 |
| **Tumor nodes** |  |  |  |  |  |  |  |  |  |  |
| 0 | 222,802 | 11,589 | 12,103 | 514 | 4.4 | 114,026 | 9,139 | 10,651 | 1,512 | 17 |
| 1 | 45,003 | 4,788 | 4,650 | -138 | -2.9 | 24,062 | 3,579 | 3,999 | 420 | 12 |
| 2-4 | 35,956 | 6,062 | 6,094 | 32 | 0.5 | 19,786 | 4,658 | 5,143 | 485 | 10 |
| 5-9 | 14,881 | 4,407 | 4,251 | -156 | -3.5 | 8,232 | 3,110 | 3,344 | 234 | 7.5 |
| 10+ | 9,231 | 4,086 | 4,030 | -56 | -1.4 | 5,088 | 2,735 | 2,863 | 128 | 4.7 |
| **Tumor grade** |  |  |  |  |  |  |  |  |  |  |
| 1 | 89,562 | 3,486 | 2,855 | -631 | -18 | 45,032 | 2,871 | 2,507 | -364 | -13 |
| 2 | 159,627 | 14,022 | 13,367 | -655 | -4.7 | 83,313 | 10,869 | 11,348 | 479 | 4.4 |
| 3 | 78,684 | 13,424 | 14,906 | 1,482 | 11 | 42,849 | 9,481 | 12,145 | 2,664 | 28 |
| **HER2 status** |  |  |  |  |  |  |  |  |  |  |
| negative | 52,502 | 4,509 | 3,718 | -791 | -18 | - | - | - | - | - |
| positive | 6,577 | 654 | 485 | -169 | -26 | - | - | - | - | - |
| **PR status** |  |  |  |  |  |  |  |  |  |  |
| negative | 49,184 | 6,837 | 5,623 | -1,214 | -18 | 26,729 | 4,651 | 4,676 | 25 | 0.5 |
| positive | 271,694 | 23,361 | 24,819 | 1,458 | 6.2 | 139,421 | 17,836 | 20,580 | 2,744 | 15 |
| **Chemotherapy** | 124,751 | 17,098 | 15,253 | -1,845 | -11 | 66,451 | 12,641 | 12,600 | -41 | -0.3 |
| **Radiotherapy** | 186,235 | 15,968 | 15,961 | -7 | 0 | 97,582 | 12,267 | 13,339 | 1,072 | 8.7 |

N: number of cases; O: observed number of deaths; P: predicted number of deaths; D: difference of the number of deaths between predicted and observed; %: percentage of the difference of the number of deaths between predicted and observed.

**Appendix 3.** Cumulative observed and predicted breast cancer-specific mortality at 10 and 15 years follow up for ER-negative patients.

| Characteristics | 10-year |  |  |  |  | 15-year |  |  |  |  |
| --- | --- | --- | --- | --- | --- | --- | --- | --- | --- | --- |
|  | N | O | P | D | % | N | O | P | D | % |
| **Total** | 82,450 | 16,599 | 19,527 | 2,928 | 18 | 47,374 | 11,045 | 13,188 | 2,143 | 19 |
| **Age at diagnosis** |  |  |  |  |  |  |  |  |  |  |
| <36y | 3,668 | 839 | 1,217 | 378 | 45 | 2,239 | 555 | 872 | 317 | 57 |
| 36-40 y | 5,369 | 1,131 | 1,431 | 300 | 26 | 3,316 | 825 | 1,049 | 224 | 27 |
| 41-50 y | 19,165 | 3,810 | 4,279 | 469 | 12 | 11,530 | 2,624 | 3,117 | 493 | 19 |
| 51-60 y | 23,735 | 4,468 | 4,938 | 470 | 11 | 13,592 | 2,987 | 3,398 | 411 | 14 |
| >60 y | 30,513 | 6,351 | 7,662 | 1,311 | 21 | 16,697 | 4,054 | 4,752 | 698 | 17 |
| **Race and ethnicity** |  |  |  |  |  |  |  |  |  |  |
| Hispanic (All Races) | 8,962 | 1,953 | 2,285 | 332 | 17 | 4,801 | 1,221 | 1,466 | 245 | 20 |
| Non-Hispanic Asian or Pacific Islander | 6,067 | 956 | 1,403 | 447 | 47 | 3,271 | 638 | 901 | 263 | 41 |
| Non-Hispanic Black | 12,548 | 3,144 | 3,231 | 87 | 2.8 | 6,903 | 1,995 | 2,121 | 126 | 6.3 |
| Non-Hispanic White | 54,873 | 10,546 | 12,608 | 2,062 | 20 | 32,399 | 7,191 | 8,700 | 1,509 | 21 |
| **Tumor size** |  |  |  |  |  |  |  |  |  |  |
| 0-10 mm | 14,252 | 1,083 | 1,221 | 138 | 13 | 7,967 | 811 | 870 | 59 | 7.3 |
| 11-20 mm | 27,611 | 3,949 | 4,707 | 758 | 19 | 16,354 | 2,783 | 3,393 | 610 | 22 |
| 21-50 mm | 33,872 | 8,602 | 10,080 | 1,478 | 17 | 19,399 | 5,691 | 6,804 | 1,113 | 20 |
| >50 mm | 6,715 | 2,965 | 3,518 | 553 | 19 | 3,654 | 1,760 | 2,121 | 361 | 21 |
| **Tumor nodes** |  |  |  |  |  |  |  |  |  |  |
| 0 | 52,840 | 6,453 | 8,126 | 1,673 | 26 | 29,602 | 4,386 | 5,544 | 1,158 | 26 |
| 1 | 10,953 | 2,552 | 2,777 | 225 | 8.8 | 6,369 | 1,655 | 1,886 | 231 | 14 |
| 2-4 | 10,102 | 3,175 | 3,667 | 492 | 16 | 6,123 | 2,134 | 2,516 | 382 | 18 |
| 5-9 | 4,866 | 2,228 | 2,486 | 258 | 12 | 3,005 | 1,456 | 1,663 | 207 | 14 |
| 10+ | 3,689 | 2,191 | 2,470 | 279 | 13 | 2,275 | 1,414 | 1,578 | 164 | 12 |
| **Tumor grade** |  |  |  |  |  |  |  |  |  |  |
| 1 | 2,475 | 214 | 257 | 43 | 20 | 1,573 | 167 | 197 | 30 | 18 |
| 2 | 15,955 | 2,704 | 2,711 | 7 | 0.3 | 9,315 | 1,880 | 1,897 | 17 | 0.9 |
| 3 | 64,020 | 13,681 | 16,559 | 2,878 | 21 | 36,486 | 8,998 | 11,094 | 2,096 | 23 |
| **HER2 status** |  |  |  |  |  |  |  |  |  |  |
| negative | 8,849 | 1,825 | 1,687 | -138 | -7.5 | - | - | - | - | - |
| positive | 2,932 | 409 | 452 | 43 | 10 | - | - | - | - | - |
| **PR status** |  |  |  |  |  |  |  |  |  |  |
| negative | 76,941 | 15,672 | 18,301 | 2,629 | 17 | 43,808 | 10,347 | 12,277 | 1,930 | 19 |
| positive | 4,903 | 833 | 1,084 | 251 | 30 | 3,116 | 614 | 793 | 179 | 29 |
| **Chemotherapy** | 56,687 | 12,087 | 13,459 | 1,372 | 11 | 31,552 | 7,896 | 8,949 | 1,053 | 13 |
| **Radiotherapy** | 43,083 | 8,424 | 9,555 | 1,131 | 13 | 25,196 | 5,646 | 6,552 | 906 | 16 |

N: number of cases; O: observed number of deaths; P: predicted number of deaths; D: difference of the number of deaths between predicted and observed; %: percentage of the difference of the number of deaths between predicted and observed.

**Appendix 4.** Cumulative observed and predicted other causes of mortality at 10 and 15 years follow up for ER-positive patients.

| Characteristics | 10-year |  |  |  |  | 15-year |  |  |  |  |
| --- | --- | --- | --- | --- | --- | --- | --- | --- | --- | --- |
|  | N | O | P | D | % | N | O | P | D | % |
| **Total** | 327,873 | 43,394 | 36,985 | -6,409 | -15 | 171,194 | 40,157 | 35,770 | -4,387 | -11 |
| **Age at diagnosis** |  |  |  |  |  |  |  |  |  |  |
| <36y | 5,925 | 93 | 76 | -17 | -18 | 3,108 | 94 | 92 | -2 | -2.4 |
| 36-40 y | 11,900 | 207 | 176 | -31 | -15 | 6,481 | 202 | 218 | 16 | 7.9 |
| 41-50 y | 63,908 | 1,541 | 1,346 | -195 | -13 | 34,024 | 1,523 | 1,620 | 97 | 6.4 |
| 51-60 y | 84,863 | 4,512 | 3,564 | -948 | -21 | 44,634 | 4,444 | 4,128 | -316 | -7.1 |
| >60 y | 161,277 | 37,041 | 31,822 | -5,219 | -14 | 82,947 | 33,894 | 29,713 | -4,181 | -12 |
| **Race and ethnicity** |  |  |  |  |  |  |  |  |  |  |
| Hispanic (All Races) | 28,532 | 2,713 | 2,421 | -292 | -11 | 13,381 | 2,303 | 2,155 | -148 | -6.4 |
| Non-Hispanic Asian or Pacific Islander | 24,937 | 1,891 | 2,113 | 222 | 12 | 12,043 | 1,711 | 1,915 | 204 | 12 |
| Non-Hispanic Black | 23,508 | 3,426 | 2,182 | -1,244 | -36 | 11,185 | 2,692 | 1,920 | -772 | -29 |
| Non-Hispanic White | 250,896 | 35,364 | 30,268 | -5,096 | -14 | 134,585 | 33,451 | 29,781 | -3,670 | -11 |
| **Tumor size** |  |  |  |  |  |  |  |  |  |  |
| 0-10 mm | 93,252 | 11,969 | 11,243 | -726 | -6.1 | 47,998 | 11,772 | 11,039 | -733 | -6.2 |
| 11-20 mm | 130,997 | 17,565 | 15,014 | -2,551 | -15 | 70,300 | 16,761 | 14,882 | -1,879 | -11 |
| 21-50 mm | 88,621 | 11,979 | 9,319 | -2,660 | -22 | 45,588 | 10,196 | 8,627 | -1,569 | -15 |
| >50 mm | 15,003 | 1,881 | 1,408 | -473 | -25 | 7,308 | 1,428 | 1,222 | -206 | -14 |
| **Tumor nodes** |  |  |  |  |  |  |  |  |  |  |
| 0 | 222,802 | 30,820 | 26,905 | -3,915 | -13 | 114,026 | 28,921 | 25,870 | -3,051 | -11 |
| 1 | 45,003 | 5,545 | 4,568 | -977 | -18 | 24,062 | 5,129 | 4,491 | -638 | -12 |
| 2-4 | 35,956 | 4,159 | 3,288 | -871 | -21 | 19,786 | 3,800 | 3,299 | -501 | -13 |
| 5-9 | 14,881 | 1,806 | 1,356 | -450 | -25 | 8,232 | 1,525 | 1,316 | -209 | -14 |
| 10+ | 9,231 | 1,064 | 867 | -197 | -18 | 5,088 | 782 | 795 | 13 | 1.6 |
| **Tumor grade** |  |  |  |  |  |  |  |  |  |  |
| 1 | 89,562 | 12,648 | 11,219 | -1,429 | -11 | 45,032 | 11,800 | 10,764 | -1,036 | -8.8 |
| 2 | 159,627 | 21,727 | 18,391 | -3,336 | -15 | 83,313 | 19,857 | 17,689 | -2,168 | -11 |
| 3 | 78,684 | 9,019 | 7,375 | -1,644 | -18 | 42,849 | 8,500 | 7,317 | -1,183 | -14 |
| **HER2 status** |  |  |  |  |  |  |  |  |  |  |
| negative | 52,502 | 6,911 | 5,629 | -1,282 | -19 | - | - | - | - | - |
| positive | 6,577 | 662 | 553 | -109 | -16 | - | - | - | - | - |
| **PR status** |  |  |  |  |  |  |  |  |  |  |
| negative | 49,184 | 6,929 | 6,004 | -925 | -13 | 26,729 | 6,603 | 6,003 | -600 | -9.1 |
| positive | 271,694 | 35,585 | 30,160 | -5,425 | -15 | 139,421 | 32,400 | 28,680 | -3,720 | -11 |
| **Chemotherapy** | 124,751 | 7,941 | 8,007 | 66 | 0.8 | 66,451 | 7,764 | 8,152 | 388 | 5 |
| **Radiotherapy** | 186,235 | 20,135 | 19,943 | -192 | -1 | 97,582 | 19,653 | 19,467 | -186 | -0.9 |

N: number of cases; O: observed number of deaths; P: predicted number of deaths; D: difference of the number of deaths between predicted and observed; %: percentage of the difference of the number of deaths between predicted and observed.

**Appendix 5.** Cumulative observed and predicted other causes of mortality at 10 and 15 years follow up for ER-positive patients.

| Characteristics | 10-year |  |  |  |  | 15-year |  |  |  |  |
| --- | --- | --- | --- | --- | --- | --- | --- | --- | --- | --- |
|  | N | O | P | D | % | N | O | P | D | % |
| **Total** | 82,450 | 8,591 | 6,717 | -1,874 | -22 | 47,374 | 8,172 | 7,003 | -1,169 | -14 |
| **Age at diagnosis** |  |  |  |  |  |  |  |  |  |  |
| <36y | 3,668 | 63 | 46 | -17 | -28 | 2,239 | 60 | 63 | 3 | 4.5 |
| 36-40 y | 5,369 | 120 | 76 | -44 | -36 | 3,316 | 121 | 106 | -15 | -12 |
| 41-50 y | 19,165 | 660 | 391 | -269 | -41 | 11,530 | 727 | 524 | -203 | -28 |
| 51-60 y | 23,735 | 1,361 | 957 | -404 | -30 | 13,592 | 1,389 | 1,184 | -205 | -15 |
| >60 y | 30,513 | 6,387 | 5,247 | -1,140 | -18 | 16,697 | 5,875 | 5,127 | -748 | -13 |
| **Race and ethnicity** |  |  |  |  |  |  |  |  |  |  |
| Hispanic (All Races) | 8,962 | 635 | 533 | -102 | -16 | 4,801 | 621 | 539 | -82 | -13 |
| Non-Hispanic Asian or Pacific Islander | 6,067 | 369 | 418 | 49 | 13 | 3,271 | 361 | 418 | 57 | 16 |
| Non-Hispanic Black | 12,548 | 1,369 | 819 | -550 | -40 | 6,903 | 1,128 | 808 | -320 | -28 |
| Non-Hispanic White | 54,873 | 6,218 | 4,948 | -1,270 | -20 | 32,399 | 6,062 | 5,238 | -824 | -14 |
| **Tumor size** |  |  |  |  |  |  |  |  |  |  |
| 0-10 mm | 14,252 | 1,516 | 1,337 | -179 | -12 | 7,967 | 1,608 | 1,391 | -217 | -14 |
| 11-20 mm | 27,611 | 3,000 | 2,352 | -648 | -22 | 16,354 | 2,988 | 2,529 | -459 | -15 |
| 21-50 mm | 33,872 | 3,446 | 2,555 | -891 | -26 | 19,399 | 3,085 | 2,622 | -463 | -15 |
| >50 mm | 6,715 | 629 | 473 | -156 | -25 | 3,654 | 491 | 462 | -29 | -5.9 |
| **Tumor nodes** |  |  |  |  |  |  |  |  |  |  |
| 0 | 52,840 | 5,864 | 4,477 | -1,387 | -24 | 29,602 | 5,704 | 4,626 | -1,078 | -19 |
| 1 | 10,953 | 1,046 | 820 | -226 | -22 | 6,369 | 979 | 864 | -115 | -12 |
| 2-4 | 10,102 | 945 | 742 | -203 | -22 | 6,123 | 860 | 790 | -70 | -8.1 |
| 5-9 | 4,866 | 431 | 392 | -39 | -9 | 3,005 | 388 | 421 | 33 | 8.4 |
| 10+ | 3,689 | 305 | 286 | -19 | -6.2 | 2,275 | 241 | 302 | 61 | 25 |
| **Tumor grade** |  |  |  |  |  |  |  |  |  |  |
| 1 | 2,475 | 317 | 282 | -35 | -11 | 1,573 | 359 | 323 | -36 | -10 |
| 2 | 15,955 | 2,022 | 1,623 | -399 | -20 | 9,315 | 1,986 | 1,708 | -278 | -14 |
| 3 | 64,020 | 6,252 | 4,813 | -1,439 | -23 | 36,486 | 5,827 | 4,973 | -854 | -15 |
| **HER2 status** |  |  |  |  |  |  |  |  |  |  |
| negative | 8,849 | 983 | 722 | -261 | -27 | - | - | - | - | - |
| positive | 2,932 | 291 | 233 | -58 | -20 | - | - | - | - | - |
| **PR status** |  |  |  |  |  |  |  |  |  |  |
| negative | 76,941 | 8,057 | 6,314 | -1,743 | -22 | 43,808 | 7,581 | 6,526 | -1,055 | -14 |
| positive | 4,903 | 459 | 350 | -109 | -24 | 3,116 | 491 | 406 | -85 | -17 |
| **Chemotherapy** | 56,687 | 3,884 | 3,553 | -331 | -8.5 | 31,552 | 3,769 | 3,675 | -94 | -2.5 |
| **Radiotherapy** | 43,083 | 3,590 | 3,325 | -265 | -7.4 | 25,196 | 3,613 | 3,541 | -72 | -2 |

N: number of cases; O: observed number of deaths; P: predicted number of deaths; D: difference of the number of deaths between predicted and observed; %: percentage of the difference of the number of deaths between predicted and observe

**Appendix 6.**
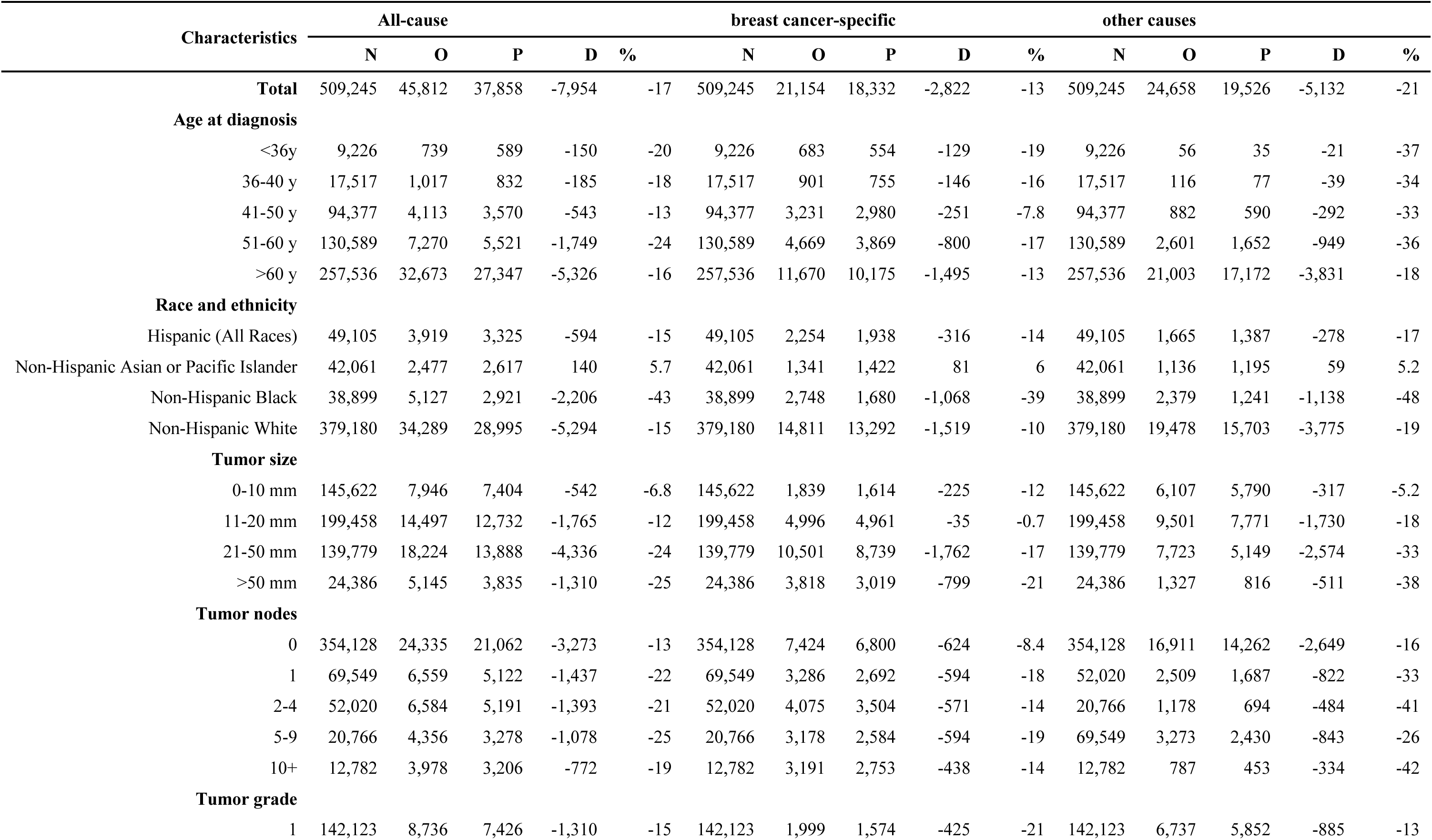
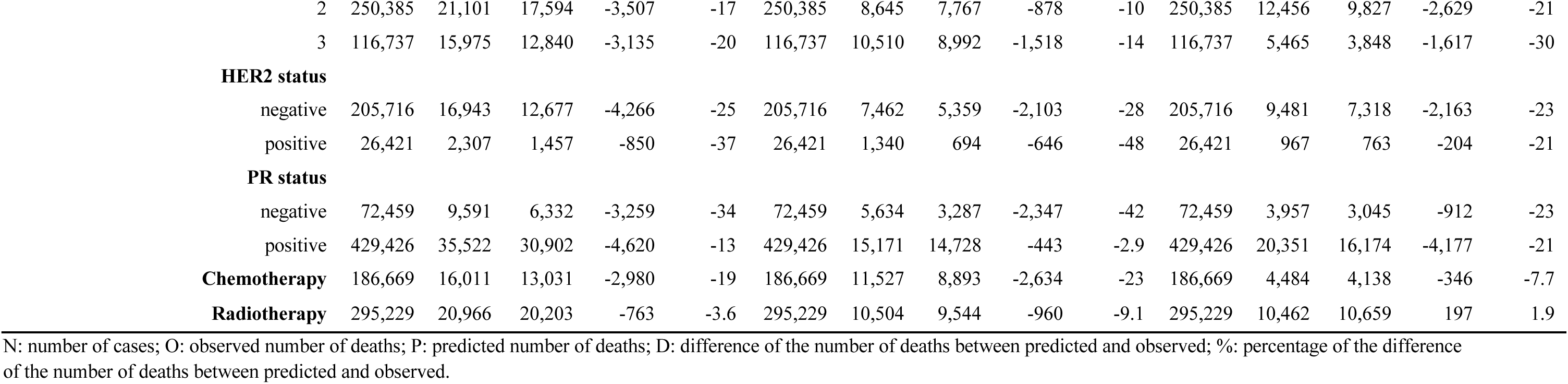
Observed and predicted all-cause, breast cancer-specific and other causes of mortality at 5 years follow up for ER-positive patients.

**Appendix 7.** Cumulative observed and predicted all-cause, breast cancer-specific and other causes of mortality at 5 years follow up for ER-negative patients.

| Characteristics | All-cause |  |  |  |  | breast cancer-specific |  |  |  |  | other causes |  |  |  |  |
| --- | --- | --- | --- | --- | --- | --- | --- | --- | --- | --- | --- | --- | --- | --- | --- |
|  | N | O | P | D | % | N | O | P | D | % | N | O | P | D | % |
| **Total** | 114,629 | 22,461 | 21,634 | -827 | -3.7 | 114,629 | 16,978 | 18,285 | 1,307 | 7.7 | 114,629 | 5,483 | 3,349 | -2,134 | -39 |
| **Age at diagnosis** |  |  |  |  |  |  |  |  |  |  |  |  |  |  |  |
| <36y | 5,087 | 953 | 1,137 | 184 | 19 | 5,087 | 898 | 1,118 | 220 | 25 | 5,087 | 55 | 19 | -36 | -66 |
| 36-40 y | 7,074 | 1,207 | 1,277 | 70 | 5.8 | 7,074 | 1,128 | 1,247 | 119 | 11 | 7,074 | 79 | 30 | -49 | -62 |
| 41-50 y | 25,289 | 4,192 | 3,846 | -346 | -8.3 | 25,289 | 3,740 | 3,689 | -51 | -1.4 | 25,289 | 452 | 157 | -295 | -65 |
| 51-60 y | 32,816 | 5,376 | 4,797 | -579 | -11 | 32,816 | 4,486 | 4,389 | -97 | -2.2 | 32,816 | 890 | 408 | -482 | -54 |
| >60 y | 44,363 | 10,733 | 10,578 | -155 | -1.4 | 44,363 | 6,726 | 7,843 | 1,117 | 17 | 44,363 | 4,007 | 2,735 | -1,272 | -32 |
| **Race and ethnicity** |  |  |  |  |  |  |  |  |  |  |  |  |  |  |  |
| Hispanic (All Races) | 13,297 | 2,542 | 2,512 | -30 | -1.2 | 13,297 | 2,118 | 2,237 | 119 | 5.6 | 13,297 | 424 | 275 | -149 | -35 |
| Non-Hispanic Asian or Pacific Islander | 9,075 | 1,215 | 1,583 | 368 | 30 | 9,075 | 964 | 1,362 | 398 | 41 | 9,075 | 251 | 221 | -30 | -12 |
| Non-Hispanic Black | 18,140 | 4,454 | 3,525 | -929 | -21 | 18,140 | 3,484 | 3,099 | -385 | -11 | 18,140 | 970 | 426 | -544 | -56 |
| Non-Hispanic White | 74,117 | 14,250 | 14,013 | -237 | -1.7 | 74,117 | 10,412 | 11,587 | 1,175 | 11 | 74,117 | 3,838 | 2,426 | -1,412 | -37 |
| **Tumor size** |  |  |  |  |  |  |  |  |  |  |  |  |  |  |  |
| 0-10 mm | 20,052 | 1,725 | 1,678 | -47 | -2.7 | 20,052 | 922 | 1,035 | 113 | 12 | 20,052 | 803 | 643 | -160 | -20 |
| 11-20 mm | 37,466 | 5,310 | 5,214 | -96 | -1.8 | 37,466 | 3,570 | 4,068 | 498 | 14 | 37,466 | 1,740 | 1,146 | -594 | -34 |
| 21-50 mm | 47,520 | 11,348 | 10,760 | -588 | -5.2 | 47,520 | 8,952 | 9,450 | 498 | 5.6 | 47,520 | 2,396 | 1,310 | -1,086 | -45 |
| >50 mm | 9,591 | 4,078 | 3,982 | -96 | -2.4 | 9,591 | 3,534 | 3,733 | 199 | 5.6 | 9,591 | 544 | 249 | -295 | -54 |
| **Tumor nodes** |  |  |  |  |  |  |  |  |  |  |  |  |  |  |  |
| 0 | 76,015 | 9,773 | 9,688 | -85 | -0.9 | 76,015 | 6,244 | 7,429 | 1,185 | 19 | 76,015 | 3,529 | 2,259 | -1,270 | -36 |
| 1 | 15,011 | 3,438 | 2,957 | -481 | -14 | 15,011 | 2,733 | 2,547 | -186 | -6.8 | 15,011 | 705 | 410 | -295 | -42 |
| 2-4 | 12,951 | 3,926 | 3,700 | -226 | -6 | 4,594 | 2,395 | 2,575 | 180 | 7.5 | 12,951 | 648 | 355 | -293 | -45 |
| 5-9 | 6,058 | 2,666 | 2,576 | -90 | -3.4 | 12,951 | 3,278 | 3,345 | 67 | 2.1 | 6,058 | 338 | 186 | -152 | -45 |
| 10+ | 4,594 | 2,658 | 2,714 | 56 | 2.1 | 6,058 | 2,328 | 2,390 | 62 | 2.7 | 4,594 | 263 | 139 | -124 | -47 |
| **Tumor grade** |  |  |  |  |  |  |  |  |  |  |  |  |  |  |  |
| 1 | 3,132 | 297 | 337 | 40 | 13 | 3,132 | 155 | 213 | 58 | 37 | 3,132 | 142 | 124 | -18 | -13 |
|  | N | O | P | D | % | N | O | P | D | % | N | O | P | D | % |
| 2 | 22,172 | 3,643 | 3,262 | -381 | -10 | 22,172 | 2,440 | 2,463 | 23 | 1 | 22,172 | 1,203 | 799 | -404 | -34 |
| 3 | 89,325 | 18,521 | 18,035 | -486 | -2.6 | 89,325 | 14,383 | 15,609 | 1,226 | 8.5 | 89,325 | 4,138 | 2,426 | -1,712 | -41 |
| **HER2 status** |  |  |  |  |  |  |  |  |  |  |  |  |  |  |  |
| negative | 31,183 | 6,373 | 4,756 | -1,617 | -25 | 31,183 | 4,761 | 3,859 | -902 | -19 | 31,183 | 1,612 | 897 | -715 | -44 |
| positive | 11,121 | 1,453 | 1,346 | -107 | -7.4 | 11,121 | 1,002 | 1,042 | 40 | 4 | 11,121 | 451 | 304 | -147 | -33 |
| **PR status** |  |  |  |  |  |  |  |  |  |  |  |  |  |  |  |
| negative | 107,058 | 21,182 | 20,302 | -880 | -4.2 | 107,058 | 16,035 | 17,153 | 1,118 | 7 | 107,058 | 5,147 | 3,149 | -1,998 | -39 |
| positive | 6,922 | 1,165 | 1,198 | 33 | 2.8 | 6,922 | 868 | 1,019 | 151 | 17 | 6,922 | 297 | 179 | -118 | -40 |
| **Chemotherapy** | 32,940 | 7,268 | 7,096 | -172 | -2.4 | 81,689 | 12,660 | 12,711 | 51 | 0.4 | 81,689 | 2,533 | 1,827 | -706 | -28 |
| **Radiotherapy** | 60,903 | 10,862 | 10,708 | -154 | -1.4 | 60,903 | 8,705 | 9,009 | 304 | 3.5 | 60,903 | 2,157 | 1,699 | -458 | -21 |

N: number of cases; O: observed number of deaths; P: predicted number of deaths; D: difference of the number of deaths between predicted and observed; %: percentage of the difference of the number of deaths between predicted and observed.

**Appendix 8.**
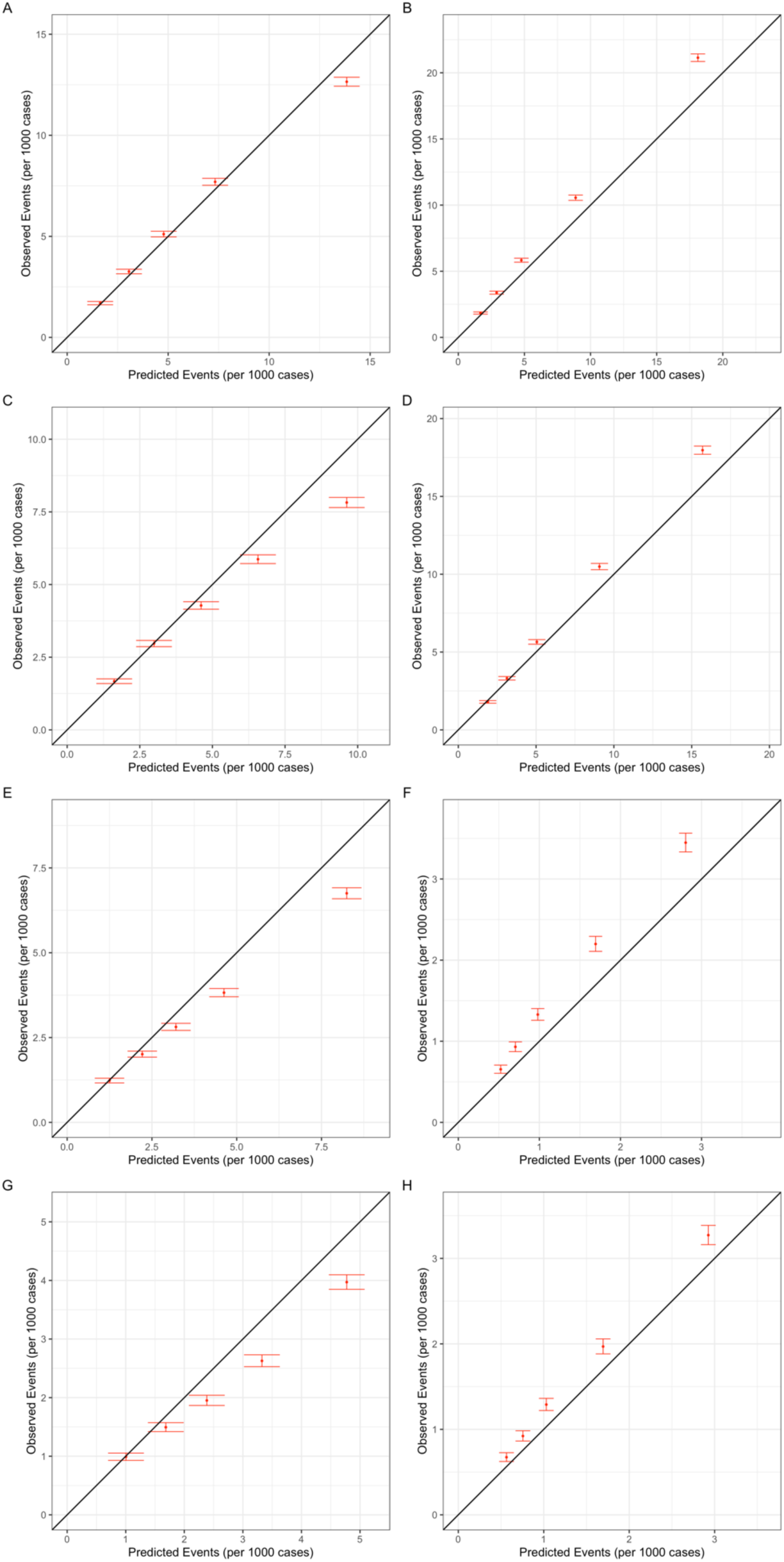
Calibration plots for (A) 10-year and (B) 15-year breast cancer-specific mortality and (C) 10-year and (D) 15-year other mortality for ER-positive breast cancer, as well as (E) 10-year and (F) 15-year breast cancer-specific mortality and (G) 10-year and (H) 15-year other mortality for ER-positive breast cancer.

**Appendix 9.**
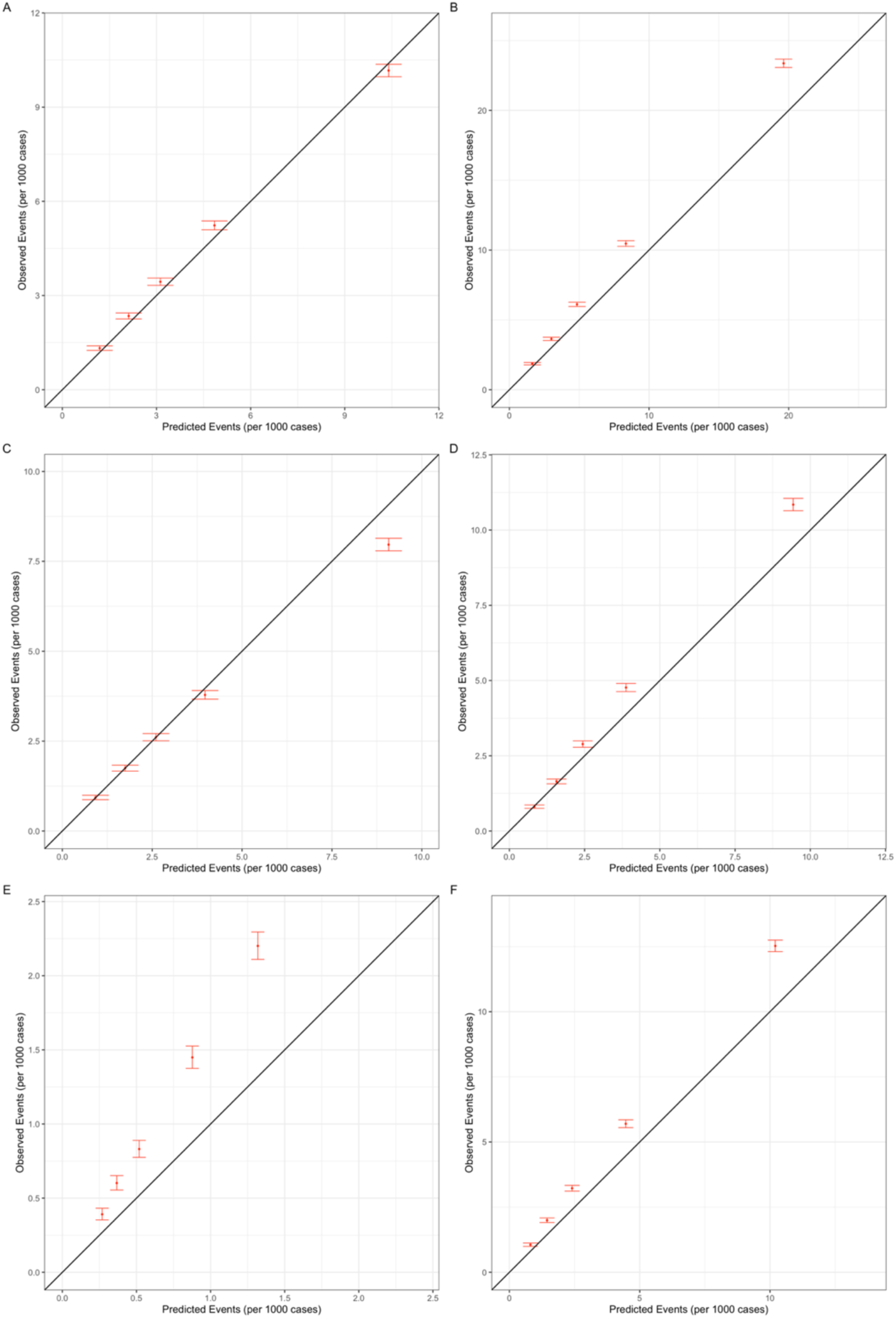
Calibration plots for 5-year mortality in ER-positive breast cancer: all-cause mortality for (A) ER-positive breast cancer and (B) ER-negative breast cancer, breast cancer-specific mortality for (C) ER-positive breast cancer and (D) ER-negative breast cancer, and mortality from other causes for (E) ER-positive breast cancer and (F) ER-negative breast cancer.

**Appendix 10.** Model discrimination (area under receiver operator characteristic curve) for 10-year and 15-year breast cancer-specific and other cause mortality by race and tumor ER-status.

| Race | ER-positive |  | ER-negative |  |
| --- | --- | --- | --- | --- |
|  | 10-year | 15-year | 10-year | 15-year |
| **Breast cancer specific mortality** |  |  |  |  |
| All | 0.768 | 0.745 | 0.73 | 0.714 |
| Non-Hispanic White | 0.767 | 0.742 | 0.722 | 0.711 |
| Non-Hispanic Asian | 0.775 | 0.752 | 0.725 | 0.714 |
| Hispanic (All Races) | 0.771 | 0.747 | 0.737 | 0.709 |
| Non-Hispanic Black | 0.753 | 0.74 | 0.73 | 0.713 |
| **Other cause mortality** |  |  |  |  |
| All | 0.794 | 0.817 | 0.769 | 0.782 |
| Non-Hispanic White | 0.795 | 0.819 | 0.73 | 0.766 |
| Non-Hispanic Asian | 0.804 | 0.818 | 0.777 | 0.788 |
| Hispanic (All Races) | 0.792 | 0.813 | 0.801 | 0.74 |
| Non-Hispanic Black | 0.752 | 0.773 | 0.773 | 0.805 |

**Appendix 11.** The discrimination for 5-year all-causes, breast cancer-specific and other causes of mortality by race and tumor ER-status.

| Race | ER-positive |  |  | ER-negative |  |  |
| --- | --- | --- | --- | --- | --- | --- |
|  | All-causes | Breast cancer-specific | Other causes | All-causes | Breast cancer-specific | Other causes |
| All | 0.758 | 0.797 | 0.763 | 0.742 | 0.751 | 0.739 |
| Non-Hispanic White | 0.76 | 0.795 | 0.764 | 0.734 | 0.739 | 0.696 |
| Non-Hispanic Asian | 0.75 | 0.793 | 0.76 | 0.737 | 0.744 | 0.755 |
| Hispanic (All Races) | 0.772 | 0.819 | 0.782 | 0.744 | 0.754 | 0.743 |
| Non-Hispanic Black | 0.736 | 0.784 | 0.734 | 0.755 | 0.763 | 0.758 |

